# A 3’UTR Insertion Is a Candidate Causal Variant at the *TMEM106B* Locus Associated with Increased Risk for FTLD-TDP

**DOI:** 10.1101/2023.07.06.23292312

**Authors:** Augustine Chemparathy, Yann Le Guen, Yi Zeng, John Gorzynski, Tanner Jensen, Chengran Yang, Nandita Kasireddy, Lia Talozzi, Michael E. Belloy, Ilaria Stewart, Aaron D. Gitler, Anthony D. Wagner, Elizabeth Mormino, Victor W. Henderson, Tony Wyss-Coray, Euan Ashley, Carlos Cruchaga, Michael D. Greicius

## Abstract

**Background and Objectives:** Single nucleotide variants near *TMEM106B* associate with risk of frontotemporal lobar dementia with TDP-43 inclusions (FTLD-TDP) and Alzheimer’s disease (AD) in genome-wide association studies (GWAS), but the causal variant at this locus remains unclear. Here we asked whether a novel structural variant on *TMEM106B* is the causal variant.

**Methods:** An exploratory analysis identified structural variants on neurodegeneration-related genes. Subsequent analyses focused on an *Alu* element insertion on the 3’UTR of *TMEM106B*. This study included data from longitudinal aging and neurogenerative disease cohorts at Stanford University, case-control cohorts in the Alzheimer’s Disease Sequencing Project (ADSP), and expression and proteomics data from Washington University in St. Louis (WUSTL). 432 individuals from two Stanford aging cohorts were whole-genome long-read and short-read sequenced. 16,906 samples from ADSP were short-read sequenced. Genotypes, transcriptomics, and proteomics data were available in 1,979 participants from an aging and dementia cohort at WUSTL. Selection criteria were specific to each cohort. In primary analyses, the linkage disequilibrium between the *TMEM106B* locus variants in the FTLD-TDP GWAS and the 3’UTR insertion was estimated. We then estimated linkage by ancestry in the ADSP and evaluated the effect of the *TMEM106B* lead variant on mRNA and protein levels.

**Results:** The primary analysis included 432 participants (52.5% females, age range 45-92 years old). We identified a 316 bp *Alu* insertion overlapping the *TMEM106B* 3’UTR tightly linked with top GWAS variants rs3173615(C) and rs1990622(A). In ADSP European-ancestry participants, this insertion is in equivalent linkage with rs1990622(A) (R^2^=0.962, D’=0.998) and rs3173615(C) (R^2^=0.960, D’=0.996). In African-ancestry participants, the insertion is in stronger linkage with rs1990622(A) (R^2^=0.992, D’=0.998) than with rs3173615(C) (R^2^=0.811, D’=0.994). In public datasets, rs1990622 was consistently associated with TMEM106B protein levels but not with mRNA expression. In the WUSTL dataset, rs1990622 is associated with TMEM106B protein levels in plasma and cerebrospinal fluid, but not with *TMEM106B* mRNA expression.

**Discussion:** We identified a novel *Alu* element insertion in the 3’UTR of *TMEM106B* in tight linkage with the lead FTLD-TDP risk variant. The lead variant is associated with TMEM106B protein levels, but not expression. The 3’UTR insertion is a lead candidate for the causal variant at this complex locus, pending confirmation with functional studies.

## Introduction

Genome-wide association studies (GWAS) using imputed micro-array data or short-read next-generation sequencing (NGS) have identified single nucleotide variants (SNVs) associated with neurodegenerative diseases. Variants associated with disease risk are frequently intergenic or intronic and rarely exonic. Among significant variants at a given locus, one may be causal or in linkage with a nearby genetic feature that is the true disease risk-modifying variant. Identifying the true causal variant at a GWAS locus is critical for elucidating disease pathogenesis and for developing candidate therapeutics. Structural variants (SVs) – which include large insertions, deletions, duplications, and other genomic features greater than 50 base pairs (bp) in length – are a source of genetic diversity whose impact on protein function is often readily interpretable due to their large size. SVs are challenging to identify with short-read NGS due to the typical 150 bp read length. SVs exceeding this length are detected in NGS data by analyzing paired and split-read evidence as well as changes in sequencing depth^1–3^. Emerging long-read sequencing (LRS) technology utilizes reads typically averaging 10 to 20 kilobases, enabling large SVs to be directly sequenced and correctly aligned to the genome^4^. LRS greatly enhances SV discovery over short-read NGS, identifying more than twice as many SVs as ensemble methods operating on short-read NGS data. Up to 83% of insertions identified by LRS are not detected by NGS algorithms^5^.

We performed LRS and SV calling for participants enrolled in Stanford’s Iqbal Farrukh and Asad Jamal Alzheimer’s Disease Research Center (ADRC) and the Stanford Aging and Memory Study (SAMS)^6^. We undertook an initial exploratory analysis looking for SVs overlapping exons, the 5’UTR or the 3’UTR of 4579 genes linked in Gene Ontology to neurodegenerative disorders. This analysis revealed a 316 base-pair insertion on the 3’UTR of *TMEM106B* which— given the strong risk-modifying effects of a *TMEM106B* locus against frontotemporal lobar dementia with TAR DNA-binding protein pathology (FTLD-TDP)—we explored further^7,8^.

The initial FTLD-TDP GWAS^7^ identified three significant SNVs (rs1990622, rs6966915, rs1020004) associated with reduced risk and in high linkage disequilibrium (LD) with one another, all on or near *TMEM106B*^7^. Subsequent work showed a pronounced effect on age-at-onset in *GRN* mutation carriers with autosomal dominant FTLD-TDP^9^. The mechanism by which variants at the *TMEM106B* locus affect FTLD-TDP risk remains unclear because the significant variants are intronic or intergenic^7^. The only coding SNV in LD with rs1990622 is the missense variant rs3173615, which results in a p.T185S amino acid change in exon 6 of *TMEM106B*. A cell-based assay suggested that rs3173615 may hasten protein degradation^10^. However, an *in vivo* study using a *GRN*^−/−^ mouse model homozygous for *TMEM106B\**S186 (the conserved residue in mice) revealed no change in TMEM106B protein levels relative to wild-type, nor amelioration of the pathological lysosomal phenotype^11^. Furthermore, a meta-analysis of FTLD-TDP GWAS evaluating both rs1990622 and rs3173615 found that rs1990622 was the most significant SNV at the locus^12^. As such, the evidence for rs3173615 as the causative variant on *TMEM106B* remains mixed. Other studies have nominated an intergenic variant near rs1990622 that alters chromatin architecture and modulates *TMEM106B* expression^13^ or 3’UTR variants that affect microRNA binding sites^14^. Recently, a large Alzheimer’s disease (AD) GWAS identified what appears to be the same *TMEM106B* locus. As in FTLD-TDP, the minor allele in Europeans was associated with reduced risk for AD, though the effect size was much smaller. Given the uncertainty regarding the causal variant at this locus, we examined whether this newly identified *TMEM106B* 3’UTR insertion may mediate the observed effect on FTLD-TDP and AD risk.

## Methods

### PARTICIPANTS AND SOURCES OF DATA

The Stanford ADRC is a cohort of healthy older controls and patients with AD and related neurological disorders (n=323 with LRS and short-read NGS, age range 45-92 years old, 169 females and 154 males, healthy controls = 150, mild cognitive impairment individuals = 60, AD cases = 30, other diagnoses = 83). The Stanford Aging and Memory Study (SAMS) is a cohort of cognitively unimpaired older individuals (n=109 with LRS and short-read NGS, age range 60-88 years old, 58 females and 51 males). The WUSTL participants were recruited as part of the Knight-ADRC and included longitudinally assessed community-dwelling adults enrolled via prospective studies of memory and aging (n=1979 with short-read NGS, transcriptomics and/or CSF and/or plasma proteomics, age range 18-103 years old, 1034 females and 945 males, healthy controls = 1005, AD cases = 858, other diagnoses = 116). Details on eligibility and assessments of participants in all datasets are provided in **eMethods**.

### LONG-READ SEQUENCING, ALIGNMENT, AND SV CALLING

High molecular weight DNA was extracted from primary blood mononuclear cells stored at −80C using a Puregene kit (Qiagen, Germany). DNA was sheared using a G-tube (Covaris LLC, Massachusetts). Sequencing libraries were prepared using Nanopore LSK-110 and sequenced on the PromethION48 (Oxford Nanopore Technologies, United Kingdom). An average of 50.4 gigabases were sequenced per sample, with a read length N50 of 18 kb. Sequencing data were base called using Guppy (High Accuracy, version 6.3), and aligned to hg38 using Minimap2^15^. Structural variants were called using Sniffles2^16^ in population mode. SVs overlapping exons, the 5’UTR or the 3’UTR of 4579 genes linked in Gene Ontology to neurodegenerative disorders and extracted using the following keywords: [“neuro”, “alzheimer”, “apolipo”, “amyloid”].

### SHORT-READ NEXT GENERATION SEQUENCING

*TMEM106B* SNV genotypes were determined from short-read NGS performed at either the Beijing Genomics Institute (BGI) in Shenzhen, China on DNBseq platform (T10 and T7), or as part of the Stanford Extreme Phenotypes in Alzheimer’s Disease project with sequencing performed at the Uniformed Services University of the Health Sciences (USUHS) on an Illumina HiSeq platform. Among the 432 participants, 29 participants were sequenced via USUHS and 403 via BGI. The whole-genome targeted coverage for both platforms was 30x and the read length was 150 bp. The Genome Analysis Toolkit (GATK) workflow germline short variant discovery was used to map genome sequencing data to the reference genome (hg38) and to produce high-confidence variant calls using joint-calling^17^.

### 3’UTR INSERTION AND SNV CALLS VALIDATION WITH IGV

The genotypes of rs1990622, rs3173615, and the 3’UTR insertion were extracted for participants with both LRS and short-read NGS available. 18 individuals with discordant doses of the three variants in LRS – where the dose of any of the three variants differed from any other – were identified for validation with IGV. The manual validation protocol is described in **eMethods**.

### COLOCALIZATION ANALYSIS

Colocalization was performed using the R package *coloc*^18^. We report the posterior probability of colocalization (PP4) between AD^19^ and FTLD-TDP^7^. Colocalization between the two GWAS results was visualized with *locuscompareR*^20^. Similarly, *coloc* analysis was performed between the plasma pQTL GWAS ^21^ ^22^ and the two neurodegenerative diseases^19,7^.

### ADSP LINKAGE DISEQUILIBRIUM ANALYSIS

Alzheimer’s disease sequencing project (ADSP) R.3 SNVs, Manta^1^, and Biograph^23^ SV calls were downloaded from NIAGADS (https://dss.niagads.org/datasets/ng00067/#data-releases). SNVs were subset to rs3173615 (7:12229791:C:G) and rs1990622 (7:12244161:A:G) using Plink 1.9^24^. SNV genotyping and SV genotypes were available in 16,906 samples. After detailed curation of the SV genotypes (**eMethods**), 16,582 unique participants in ADSP had a robust calling of the 3’UTR insertion.

To identify European and African ancestry individuals in the ADSP, the ancestries of all ADSP individuals were determined using SNPWeights v2^25^ with reference populations from the 1000 Genomes Consortium^26^. Individuals with greater than 75% African global ancestry were classified as African ancestry^27^, and similarly for European ancestry^28^.

### eQTL AND pQTL ASSOCIATIONS WITH THE *TMEM106B* Locus

Expression quantitative trait locus (eQTL) and protein quantitative trait locus (pQTL) effect sizes and p-values were queried for rs1990622 from summary statistics (GTEx^29^, MetaBrain^30^, eQTLGen^31^ for eQTLs and ARIC^22^, DECODE^32^, Wingo^33^ for pQTLs).

Additionally, we analyzed a novel dataset collected at WUSTL, which included 1,979 participants with NGS (1,979 participants), blood bulk RNASeq (428 European participants, all healthy controls), plasma (1,150 European and 200 African participants, with 711 European and 120 African healthy controls), and CSF (1,210 European participants, with 588 healthy controls) proteomics obtained using SomaScan aptamers. Global ancestry was estimated using principal component analysis using the 1000 Genomes ancestry as anchors, and individuals were separated into European and African ancestry groups (**eTable 1** provides the demographics by analyses). Details on the ‘omics analysis methods are provided in **eMethods**. These were adjusted for age and sex, due to the association of TMEM106B and GRN protein levels with these two demographic factors (**eFigures 1-3**).

#### Standard Protocol Approvals, Registrations, and Patient Consents

Participants or their caregivers provided written informed consent in the original studies. Study protocols were approved by the Institutional Review Boards at Stanford University and Washington University in St. Louis (WUSTL).

#### Data Availability

Data included in this paper will be provided in anonymized form upon reasonable request made via email to the corresponding author and completion of a Material Transfer Agreement.

## RESULTS

We carried out whole-genome LRS and SV calling for 432 participants enrolled in the Stanford ADRC or SAMS. In total, we identified 208 SVs overlapping an exon, 3’UTR, or 5’UTR site of one of the 4579 genes considered. Among these, we identified a common 322 bp deletion on the 3’UTR of *TMEM106B* for further analysis given the interest in this locus (**Figure 1a)**^34^. This SV overlaps a 316 bp *Alu*Yb8 mobile element that is prevalent in European-ancestry individuals and is included in the hg38 reference genome^35^. Our analysis pipeline therefore detected this SV as a 322 bp deletion when comparing individual genomes to the hg38 reference. We will refer to the SV, hereafter, as an insertion.

**Figure 1.**
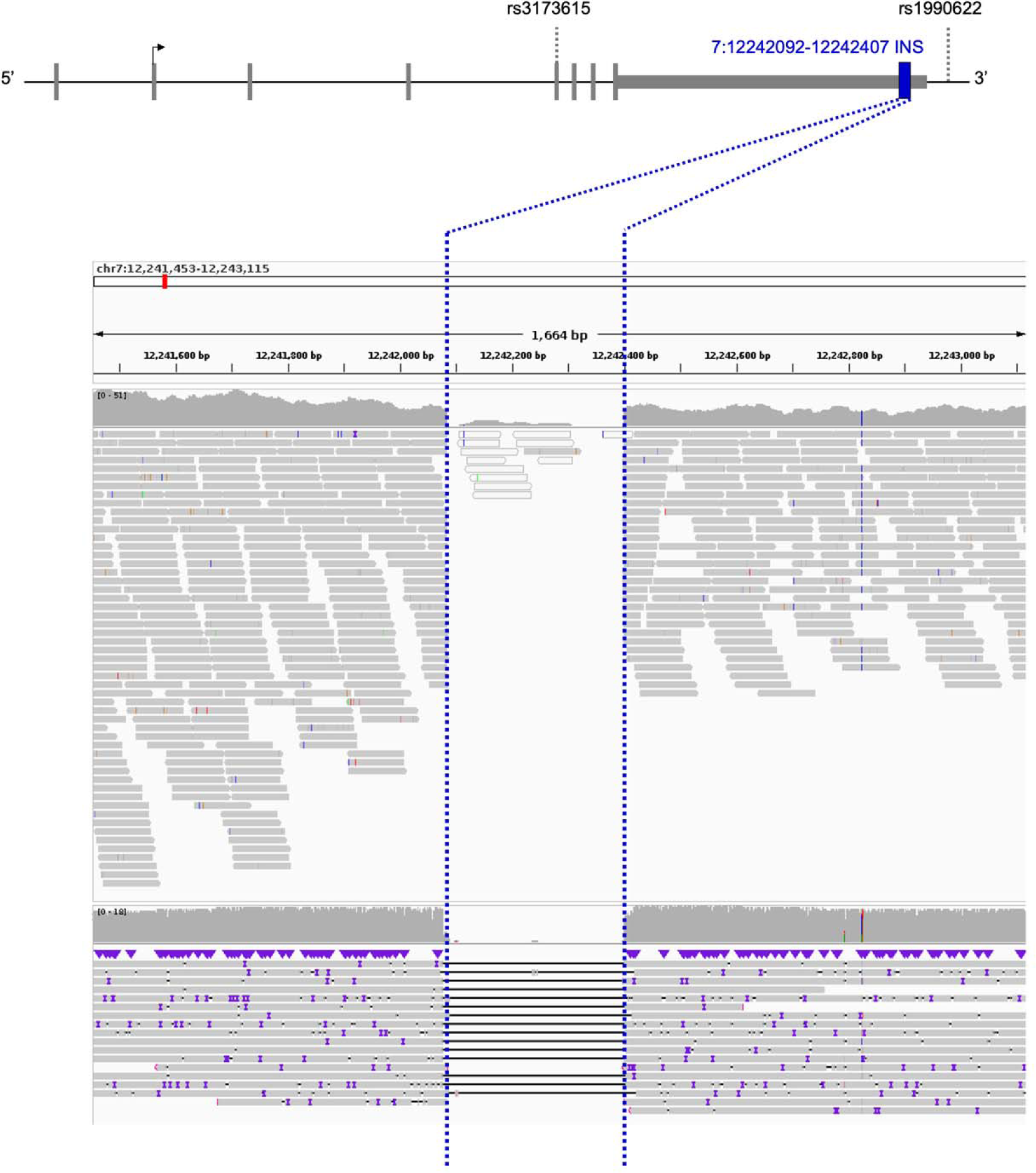
*TMEM106B* locus landscape and 3’ UTR insertion location. (Top) *TMEM106B* 3’ UTR insertion position relative to SNVs associated with FTLD-TDP. (Bottom) The *TMEM106B* 3’ UTR insertion, detected as a deletion compared to the reference genome, was seen in both next-generation sequencing (upper panel) and long-read sequencing (lower panel). The region corresponding to the structural variant is delineated with blue dotted lines. Both sequencing modalities are displayed here for a representative cohort participant without the *TMEM106B* 3’ UTR insertion.

The insertion is highly prevalent with allele frequency (AF)=0.535 in our LRS dataset, comparable to the major allele frequency, in European non-Finnish ancestry, in gnomAD (v3.1.2) of rs1990622(A) (AF=0.586) and rs3173615(C) (AF=0.587). The insertion was detectable in both LRS and short-read NGS (**Figure 1b**). We linked LRS data to high coverage short-read NGS data to evaluate the LD between the 3’UTR insertion, rs1990622(A) and rs3173615(C). The *TMEM106B* 3’UTR insertion was in perfect LD with rs1990622(A) and rs3173615(C) in all but two Stanford individuals. For the sake of consistency, we will display all SNV data with the risk allele that is most often co-inherited with the insertion. This is the A allele for rs1990622 and the C allele for rs3173615. Note that the insertion, rs1990722(A) and rs3173615(C) are the major alleles in European-ancestry individuals (AF ∼0.59) but the minor alleles in African-ancestry individuals (AF ∼0.27).

The linkage between the *TMEM106B* 3’UTR insertion, rs1990622(A), and rs3173615(C) was then established in a large cohort by querying the ADSP database. 10,265 European ancestry and 2,212 African ancestry participants’ samples were genotyped at both SNVs and the SV, with respective AFs for the *Alu* insertion of AF_EUR_=0.591 and AF_AFR_=0.272. In European ancestry individuals, The *Alu* insertion is in comparable LD with rs1990622 (R^2^=0.962, D’=0.998) and rs3173615 (R^2^=0.960, D’=0.996). In African ancestry individuals in ADSP, the *Alu* insertion is in stronger LD with rs1990622 (R^2^=0.992, D’=0.998) than with rs3173615 (R^2^=0.811, D’=0.994).

A colocalization analysis (**Figure 2**) demonstrated that the signals identified in the FTLD-TDP GWAS^7^and AD GWAS ^19^ at the *TMEM106B* locus have the same linkage structure (PP4 = 99.5%), suggesting that the same genetic signal is driving effects across both disorders.

**Figure 2.**
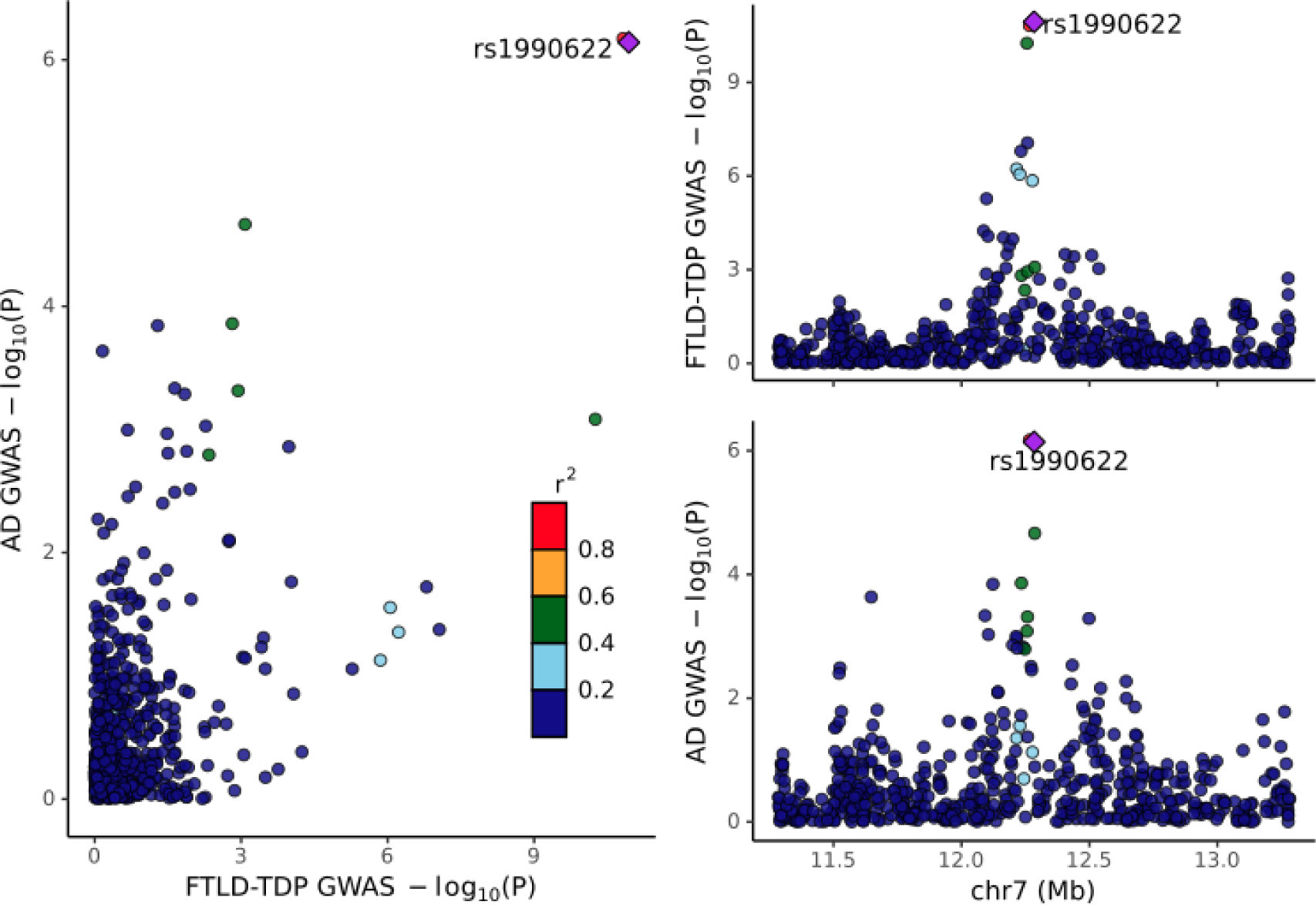
Colocalization between the AD^19^ and FTLD-TDP^7^ genome-wide association studies. The posterior probability of colocalization (PP4) = 99.5%. Note that the FTLD-TDP GWAS only included directly genotyped variants, i.e., not imputed, and thus the intersection of the two GWAS on this 2MB window is somewhat sparse, including only 607 variants.

We next evaluated the effect of rs1990622 on *TMEM106B* expression and protein levels in eQTL and pQTL datasets (**Table 1**). In a large brain eQTL dataset (European ancestry - Metabrain; n=6,601), rs1990622(A) was not associated with *TMEM106B* expression. In a smaller brain eQTL dataset (African ancestry - Metabrain; n=1,016), rs1990622(A) was significantly associated with increased *TMEM106B* expression. In two plasma eQTL datasets (GTEx, n=670; and eQTLGen, n=31,247), rs1990622(A) was significantly associated with decreased *TMEM106B* expression. In three large plasma pQTL datasets (deCODE; n=35,371; ARIC European; n=7,213; ARIC African; n=1,871), rs1990622(A) was significantly associated with increased TMEM106B protein levels. In a smaller brain pQTL dataset (Wingo; n=722), rs1990622(A) was not significantly associated with TMEM106B protein levels (p=0.1) but the direction of the effect was consistent with the plasma pQTL results. In addition, colocalization analyses showed that the plasma-based pQTL association with TMEM106B level in deCODE and ARIC colocalizes with the signal in AD and FTLD-TDP at the *TMEM106B* locus. Notably, all posterior probabilities of colocalization between the densely imputed AD GWAS and TMEM106B protein level associations in plasma were above 80% (PP4 with deCODE = 81.1%; PP4 with ARIC European = 86.3%; PP4 with ARIC African = 91.6%) (**eFigure 4**).

**Table 1.** Effect of rs1990622 as a *TMEM106B* expression quantitative trait locus (eQTL) or protein quantitative trait locus (pQTL). All effect sizes are reported for the tested A allele with the G allele set as reference at rs1990622. Parameter estimates are those reported by respective studies and may be on different scales given different normalization procedures for transcriptomic and proteomic data. *Negative Z-value indicating that A allele is associated with decreased *TMEM106B* expression. Abbreviations: EA: European Ancestry, AA: African Ancestry.

| Dataset | Tissue | Sample size | Effect size | p-value |
| --- | --- | --- | --- | --- |
| **Expression (e)QTL** |  |  |  |  |
| eQTLGen | Blood | 31,427 | - * | 0.0174 |
| GTEx | Blood | 670 | -0.25 | 4.00E-30 |
| Metabrain EA | Brain (cortex) | 6,601 | +0.0268 | 0.3115 |
| Metabrain AA | Brain (cortex) | 1,016 | +0.2053 | 0.0031 |
| **Protein (p)QTL** |  |  |  |  |
| deCODE | Plasma | 35,371 | +0.1106 | 3.69E-43 |
| ARIC EA | Plasma | 7,213 | +0.2547 | 7.33E-53 |
| ARIC AA | Plasma | 1,871 | +0.3147 | 1.50E-19 |
| Wingo | Brain (cortex) | 722 | +0.0773 | 0.1005 |

To expand beyond these eQTL and pQTL results from publicly available databases, we tested the association between rs1990622 and *TMEM106B* expression, TMEM106B protein levels, and GRN protein levels in the WUSTL Neurogenomics dataset^36,37,38^. First, we examined GRN and TMEM106B protein levels in a group of 11 carriers of FTLD-TDP-causing progranulin (*PGRN*) mutations. The *PGRN* mutation carriers have significantly higher plasma levels of TMEM106B compared to non-carriers from the same families (p < 0.05, **Figure 3a**). GRN levels were significantly lower in *PRGN* mutation carriers compared to non-carriers from the same families (p < 0.001, **Figure 3b**). With only 11 *PGRN* mutation carriers we could not determine if the *TMEM106B* locus variants impacted TMEM106B or GRN levels. When comparing AD cases versus older controls we found that plasma TMEM106B levels were significantly lower in AD (β = −0.011; 95% CI [−0.019; −0.004]; p = 4.25×10^−3^) as were CSF TMEM106B levels (β = −0.186; 95% CI [−0.277; −0.095]; p = 6.35×10^−5^). We also found a significant increase in plasma (β = 0.016; 95% CI [0.002; 0.031]; p =0.021) and CSF GRN levels (β = 0.0969; 95% CI [0.004; 0.19]; p = 0.04) when comparing AD cases with controls.

**Figure 3.**
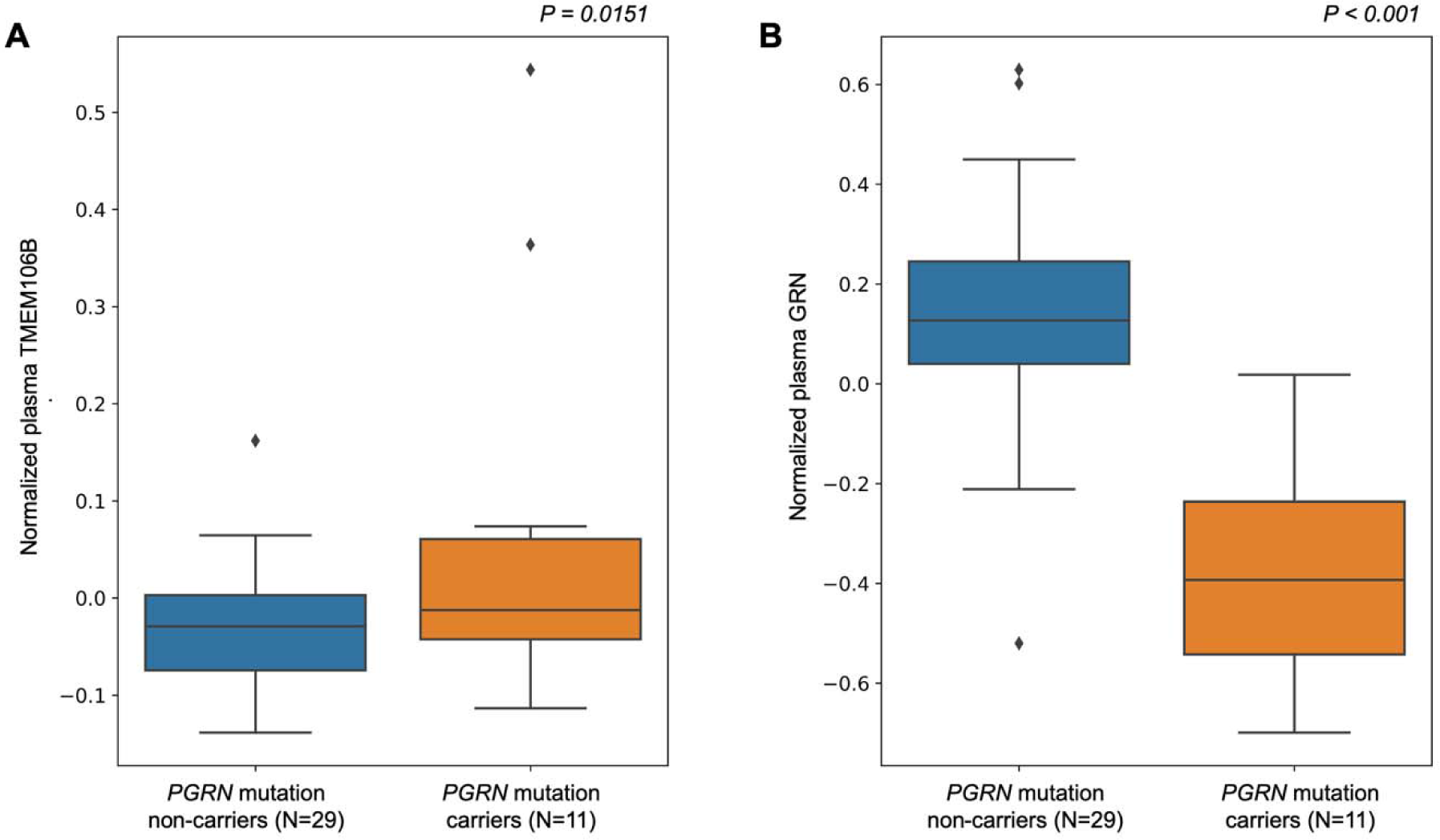
Differential abundance analysis of *PGRN* mutation carriers versus non-carriers on plasma TMEM106B and GRN protein levels. A) Boxplot of the normalized plasma protein *TMEM106B* levels between *PGRN* mutation carriers and non-carriers. Mutation carriers have significantly higher TMEM106B levels (p = 0.0151). B) Boxplot of normalized plasma protein *GRN* levels between *PGRN* mutation carriers and non-carriers. Mutation carriers have significantly lower GRN levels (p = 2.89×10^−8^), Analyses were adjusted for age, sex, and proteomic principal components 1 and 2.

In the large WUSTL cohort of healthy older European-ancestry controls, rs1990622 was not associated with *TMEM106B* expression levels (p = 0.87) in blood but was associated with TMEM106B protein levels in both plasma and CSF (**Figure 4)**. rs1990622(A) was significantly associated with increased levels of TMEM106B in plasma (β = 0.0227; 95% CI [0.015;0.031]; p = 5.6×10^−8^) and in CSF (β = 0.280; 95% CI [0.232;0.327]; p = 3.9×10^−27^), with a roughly 10-fold larger effect size in CSF. rs1990622(A) was also associated with TMEM106B protein levels in plasma of African-ancestry controls (β = 0.0290; 95% CI [0.002;0.057]; p = 0.0387 and **eTable 2**). There were not enough African-ancestry CSF samples to perform similar analyses. In examining GRN levels, we did not find a significant association of rs1990622(A) with GRN levels in plasma or CSF.

**Figure 4.**
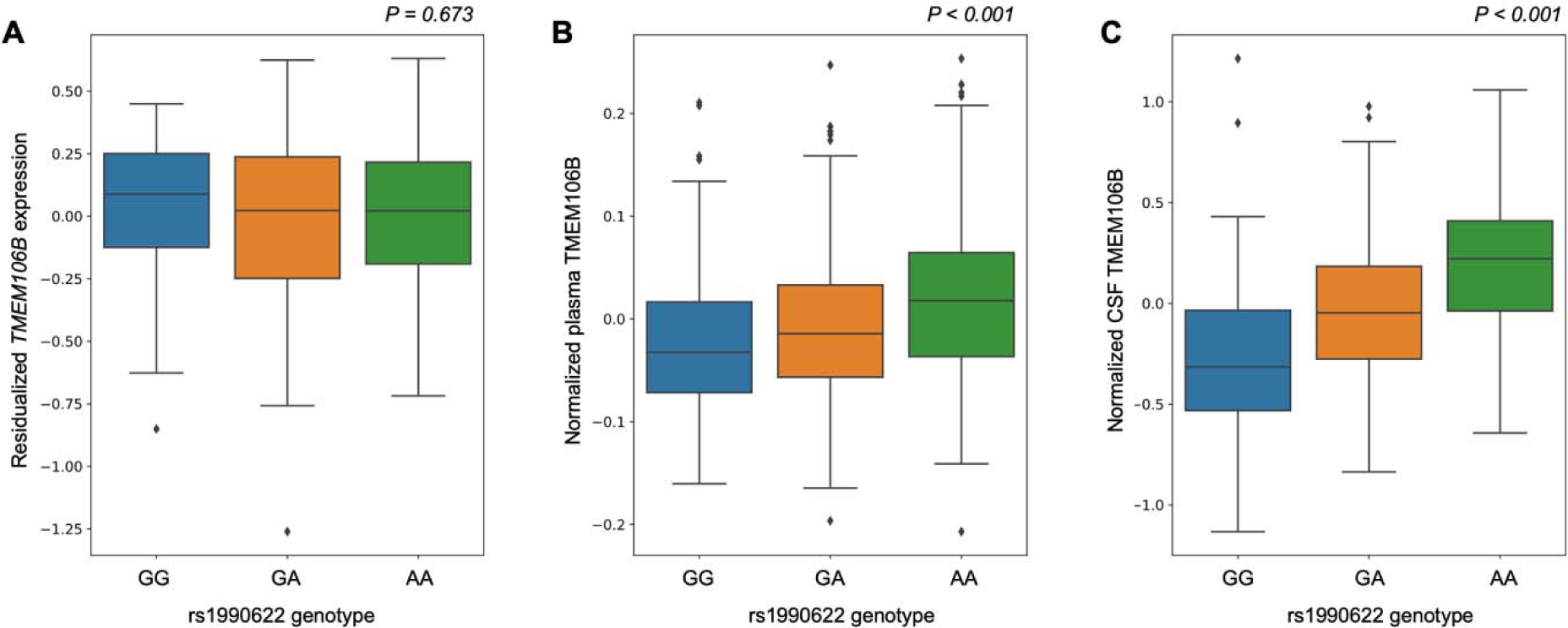
Effects of rs1990622 on *TMEM106B* gene expression in blood and TMEM106B protein levels in plasma and cerebrospinal fluid of European-ancestry controls. A) Boxplot of the residualized *TMEM106B* gene expression in healthy older control blood samples across the rs1990622 genotypes (beta = 0.00481, p = 0.673). B) Boxplot of normalized TMEM106B protein levels from plasma in healthy older controls across the rs1990622 genotypes (beta = 0.0215, p = 6.3×10^−11^). C) Boxplot of normalized TMEM106B protein levels from cerebrospinal fluid in healthy older controls across the rs1990622 genotypes (beta = 0.283, p = 2.5×10^−32^). All analyses were adjusted for age and sex.

Since rs1990622 and rs3173615 were called directly from NGS in the WUSTL data we could compare the effect sizes of the two SNVs directly. Results were nearly identical with a numerically larger effect of rs1990622 in plasma and CSF of European-ancestry subjects and a numerically larger effect of rs3173615 in plasma of African-ancestry subjects (**eTable 2, eFigure 5**).

## DISCUSSION

The *TMEM106B* 3’UTR *Alu*Yb8 insertion is a previously unreported variant that may mediate the effect of the *TMEM106B* locus on FTLD-TDP risk and AD risk. At present, the candidates for the causal variant at this locus are (1) the *TMEM106B* 3’UTR insertion described here; (2) rs3173615, the only coding SNV in this linkage block; (3) rs1990622, the leading GWAS hit; or (4) another variant in this linkage block.

Here we used publicly available expression and protein datasets to establish that rs1990622 is a consistent pQTL but an inconsistent eQTL and then confirmed this in a large, novel transcriptomics and proteomics dataset from WUSTL. This suggests that the effect at the *TMEM106B* locus is likely mediated by a genetic variant that acts after transcription to impact TMEM106B protein levels. Because intronic and intergenic variants are not incorporated into the processed mRNA molecule it is less likely that such variants, including rs1990622, are directly mediating the effect of this locus. Furthermore, the pQTL findings, coupled with the finding of increased TMEM106B protein levels in *PGRN* mutation carriers, suggest that the *TMEM106B* locus exerts its effect by altering protein availability rather than by altering protein function due to an amino acid change. This reduces the likelihood that rs3173615 is the causal variant. A prior *in vitro* study suggested that the TMEM106B *p.*S185 protein is less stable than wild-type and may therefore result in reduced protein levels^10^, but recent *in vivo* mouse model work suggests that the rs3173615 missense variant is not causal^11^. Moreover, rs1990622 was found to be more significant than rs3173615 in an FTLD-TDP GWAS meta-analysis^12^, which would be unexpected if the protective effect of the rs1990622 minor allele were due to its linkage with rs3173615.

We found in ADSP that the *TMEM106B* 3’UTR insertion is in higher LD with rs1990622(A) than rs3173615(C), which is consistent with a model in which the respective significance of these two SNVs is related to their linkage with the *TMEM106B* 3’UTR insertion. The proteomics data from WUSTL showed that the effect sizes of these variants are almost identical for CSF and plasma TMEM106B levels with a minimally larger effect for rs1990622 in European-ancestry subjects and a minimally larger effect for rs3173615 in African-ancestry individuals. We emphasize here that the differences between the two variants on protein levels are exceedingly small and should not be considered significant. Directly comparing the effect size of the two SNVs in the same set of subjects, would be more powerful in a large sample of African-ancestry AD cases and controls or a larger sample of African-ancestry proteomics dataset given that roughly 6% of African-ancestry haplotypes are discordant across these two SNVs compared to only ∼0.5% of European-ancestry haplotypes. With the growing focus on enrolling more underrepresented minority participants in genetics studies, such an analysis should be feasible in the near future. Given the high LD across the locus, such analyses will be more reliable and definitive when carried out on short-read NGS or LRS datasets rather than on imputed SNV array datasets.

There are several possibilities as to how the *TMEM106B* 3’UTR *Alu* insertion could increase risk in FTLD-TDP and AD. The pQTL evidence indicates that the major risk allele at the *TMEM106B* locus increases TMEM106B protein levels both in plasma and in CSF. In cortex, the trend is in the same direction but did not reach significance (p = 0.10) with this relatively smaller sample size (Table 1). In a small group of *PGRN* mutation carriers, we replicate prior work ^9,39,40^ showing reduced plasma levels of GRN and demonstrate, for the first time, that TMEM106B levels are increased. As FTLD-TDP due to *PGRN* mutations is thought to result from haploinsufficiency of GRN^34,41,42^ a compelling hypothesis is that the *Alu* insertion on the 3’UTR results in an increase of TMEM106B levels in the brain which further depletes GRN levels. These two proteins are both major components of the endolysosomal pathway and so are well-positioned to interact^37,43^ Further evidence that the two proteins interact comes from recent work demonstrating that TMEM106B fibrils form aggregates in a variety of neurodegenerative disorders and as a function of age, but that these TMEM106B fibrils are far more prominent in *PGRN* mutation carriers with FTLD-TDP^44–47^. In AD we found the opposite pattern (reduced TMEM106B levels and increased GRN levels in plasma and in CSF). This is less readily interpretable given that we do not understand GRN’s role in AD. One possibility is that the findings in AD reflect compensatory changes in the setting of pathogenesis rather than causal effects driving pathogenesis.

The mechanism by which the insertion may impact TMEM106B levels remains uncertain. The insertion may result in selective enrichment of an alternate transcript polyadenylation site, changing the 3’UTR. Such a change in the 3’UTR could affect protein binding, which may in turn impact translational efficiency, alter the subcellular localization of the RNA, or impair protein routing to the endoplasmic reticulum^48^. Identifying a clear-cut mechanism linking the insertion to increased TMEM106B protein levels is still required to confirm that this is the causal variant at the locus.

In summary, we report a 316 bp *Alu* insertion on *TMEM106B* that is in tight LD with rs1990622(A) and rs3173615(C) in a large LRS dataset. LRS provides a valuable tool for the detection of large genomic variants that can aid in the interpretation of GWAS results and elucidate genetic drivers of disease. Ongoing functional work by our group and others will elaborate on the mechanisms connecting the 3’UTR *Alu*Yb8 insertion to pathogenesis in FTLD-TDP and AD.

## Data Availability

Data will be made available after the paper is accepted by a peer-reviewed journal.

## FUNDING AND ACKNOWLEDGMENTS

This work was supported by the following NIH grants to Stanford investigators: RO1AG060747; R35AG072290; R01AG048076; R01AG074339; P30AG066515; and K99AG075238.This work was also supported by the Alzheimer’s Association (AARF-20-683984).

Multiomics data generated by the NeuroGenomics and Informatics Funding was supported by grants from the NIH:R01AG044546; P01AG003991; RF1AG053303: RF1AG058501; U01AG058922;, RF1AG074007; the Chan Zuckerberg Initiative (CZI); the Michael J. Fox Foundation; the Department of Defense (W81XWH2010849); the Alzheimer’s Association Zenith Fellows Award (ZEN-22-848604); and an Anonymous foundation. The recruitment and clinical characterization of research participants at Washington University were supported by NIH P30AG066444; P01AG03991; and P01AG026276. This work was supported by access to equipment made possible by the Hope Center for Neurological Disorders, the Neurogenomics and Informatics Center (NGI: https://neurogenomics.wustl.edu/), and the Departments of Neurology and Psychiatry at Washington University School of Medicine. The following investigators assisted in the preparation of the WUSTL ‘omics dataset Matt Johnson, Maulikkumar Patel, Maria Victoria Fernandez, Jessie Sanford, Devin Dikec, Ellen Liu, Dan Western, Thomas Marsh, Priyanka Gorijala, Jigyasha Timsina, and Judy Lihua Wang.

