## Supplemental Appendix for "A 3’UTR Insertion Is a Candidate Causal Variant at the *TMEM106B* Locus Associated with Increased Risk for FTLD-TDP"

**eAppendix.** Summary Statistics Data Availability

**eAppendix.** Additional Acknowledgments.

**eMethods.** Supplementary Note.

**eFigure 1.** Effect of age on TMEM106B protein level in CSF and plasma.

**eFigure 2.** Effect of age on TMEM106B or GRN levels in two tissue types in two ancestries

**eFigure 3**. Effect of sex on TMEM106B or GRN levels in two tissue types in two ancestries

**eFigure 4**. Colocalization analyses between the AD and FTLD-TDP GWAS and the plasma pQTL GWAS (deCODE and ARIC).

**eFigure 5.** Effects of variants rs1990622(A) and rs3173615(C) on levels of protein TMEM106B in two tissues.

**eTable 1.** Demographics per ‘omics modalities in the WUSTL Neurogenomics dataset.

**eTable 2.** Association of TMEM106B variants across tissues and in multiple populations.

**eReferences.**

**eAppendix.** Summary Statistics Data Availability

eQTLgen eQTL: [https://www.eqtlgen.org](https://www.eqtlgen.org/)

GTEx eQTL: <http://ftp.ebi.ac.uk/pub/databases/spot/eQTL/imported/GTEx_V8/>

MetaBrain eQTL: <https://www.metabrain.nl>

ARIC pQTL: <http://nilanjanchatterjeelab.org/pwas/>

DECODE pQTL: [https://www.decode.com/summarydata/](https://www.nature.com/articles/s41588-021-00978-w#MOESM4)

Wingo pQTL: <https://www.synapse.org/#!Synapse:syn31826294>

**eAppendix.** Additional Acknowledgments

Data for this study were prepared, archived, and distributed by the National Institute on Aging Alzheimer’s Disease Data Storage Site (NIAGADS) at the University of Pennsylvania (U24-AG041689), funded by the National Institute on Aging.

**Acknowledgments for the use of ADSP WGS data**

The Alzheimer’s Disease Sequencing Project (ADSP) is comprised of two Alzheimer’s Disease (AD) genetics consortia and three National Human Genome Research Institute (NHGRI) funded Large Scale Sequencing and Analysis Centers (LSAC). The two AD genetics consortia are the Alzheimer’s Disease Genetics Consortium (ADGC) funded by NIA (U01 AG032984), and the Cohorts for Heart and Aging Research in Genomic Epidemiology (CHARGE) funded by NIA (R01 AG033193), the National Heart, Lung, and Blood Institute (NHLBI), other National Institute of Health (NIH) institutes and other foreign governmental and non-governmental organizations. The Discovery Phase analysis of sequence data is supported through UF1AG047133 (to Drs. Schellenberg, Farrer, Pericak-Vance, Mayeux, and Haines); U01AG049505 to Dr. Seshadri; U01AG049506 to Dr. Boerwinkle; U01AG049507 to Dr. Wijsman; and U01AG049508 to Dr. Goate and the Discovery Extension Phase analysis is supported through U01AG052411 to Dr. Goate, U01AG052410 to Dr. Pericak-Vance and U01 AG052409 to Drs. Seshadri and Fornage.

Sequencing for the Follow Up Study (FUS) is supported through U01AG057659 (to Drs. PericakVance, Mayeux, and Vardarajan) and U01AG062943 (to Drs. Pericak-Vance and Mayeux). Data generation and harmonization in the Follow-up Phase is supported by U54AG052427 (to Drs. Schellenberg and Wang). The FUS Phase analysis of sequence data is supported through U01AG058589 (to Drs. Destefano, Boerwinkle, De Jager, Fornage, Seshadri, and Wijsman), U01AG058654 (to Drs. Haines, Bush, Farrer, Martin, and Pericak-Vance), U01AG058635 (to Dr. Goate), RF1AG058066 (to Drs. Haines, Pericak-Vance, and Scott), RF1AG057519 (to Drs. Farrer and Jun), R01AG048927 (to Dr. Farrer), and RF1AG054074 (to Drs. Pericak-Vance and Beecham).

The ADGC cohorts include: Adult Changes in Thought (ACT) (U01 AG006781, U19 AG066567), the Alzheimer’s Disease Research Centers (ADRC) (P30 AG062429, P30 AG066468, P30 AG062421, P30 AG066509, P30 AG066514, P30 AG066530, P30 AG066507, P30 AG066444, P30 AG066518, P30 AG066512, P30 AG066462, P30 AG072979, P30 AG072972, P30 AG072976, P30 AG072975, P30 AG072978, P30 AG072977, P30 AG066519, P30 AG062677, P30 AG079280, P30 AG062422, P30 AG066511, P30 AG072946, P30 AG062715, P30 AG072973, P30 AG066506, P30 AG066508, P30 AG066515, P30 AG072947, P30 AG072931, P30 AG066546, P20 AG068024, P20 AG068053, P20 AG068077, P20 AG068082, P30 AG072958, P30 AG072959), the Chicago Health and Aging Project (CHAP) (R01 AG11101, RC4 AG039085, K23 AG030944), Indiana Memory and Aging Study (IMAS) (R01 AG019771), Indianapolis Ibadan (R01 AG009956, P30 AG010133), the Memory and Aging Project (MAP) ( R01 AG17917), Mayo Clinic (MAYO) (R01 AG032990, U01 AG046139, R01 NS080820, RF1 AG051504, P50 AG016574), Mayo Parkinson’s Disease controls (NS039764, NS071674, 5RC2HG005605), University of Miami (R01 AG027944, R01 AG028786, R01 AG019085, IIRG09133827, A2011048), the Multi-Institutional Research in Alzheimer’s Genetic Epidemiology Study (MIRAGE) (R01 AG09029, R01 AG025259), the National Centralized Repository for Alzheimer’s Disease and Related Dementias (NCRAD) (U24 AG021886), the National Institute on Aging Late Onset Alzheimer’s Disease Family Study (NIA- LOAD) (U24 AG056270), the Religious Orders Study (ROS) (P30 AG10161, R01 AG15819), the Texas Alzheimer’s Research and Care Consortium (TARCC) (funded by the Darrell K Royal Texas Alzheimer’s Initiative), Vanderbilt University/Case Western Reserve University (VAN/CWRU) (R01 AG019757, R01 AG021547, R01 AG027944, R01 AG028786, P01 NS026630, and Alzheimer’s Association), the Washington Heights-Inwood Columbia Aging Project (WHICAP) (RF1 AG054023), the University of Washington Families (VA Research Merit Grant, NIA: P50AG005136, R01AG041797, NINDS: R01NS069719), the Columbia University Hispanic Estudio Familiar de Influencia Genetica de Alzheimer (EFIGA) (RF1 AG015473), the University of Toronto (UT) (funded by Wellcome Trust, Medical Research Council, Canadian Institutes of Health Research), and Genetic Differences (GD) (R01 AG007584). The CHARGE cohorts are supported in part by National Heart, Lung, and Blood Institute (NHLBI) infrastructure grant HL105756 (Psaty), RC2HL102419 (Boerwinkle) and the neurology working group is supported by the National Institute on Aging (NIA) R01 grant AG033193.

The CHARGE cohorts participating in the ADSP include the following: Austrian Stroke Prevention Study (ASPS), ASPS-Family study, and the Prospective Dementia Registry-Austria (ASPS/PRODEM-Aus), the Atherosclerosis Risk in Communities (ARIC) Study, the Cardiovascular Health Study (CHS), the Erasmus Rucphen Family Study (ERF), the Framingham Heart Study (FHS), and the Rotterdam Study (RS). ASPS is funded by the Austrian Science Fond (FWF) grant number P20545-P05 and P13180 and the Medical University of Graz. The ASPS-Fam is funded by the Austrian Science Fund (FWF) project I904), the EU Joint Programme – Neurodegenerative Disease Research (JPND) in frame of the BRIDGET project (Austria, Ministry of Science) and the Medical University of Graz and the Steiermärkische Krankenanstalten Gesellschaft. PRODEM-Austria is supported by the Austrian Research Promotion agency (FFG) (Project No. 827462) and by the Austrian National Bank (Anniversary Fund, project 15435. ARIC research is carried out as a collaborative study supported by NHLBI contracts (HHSN268201100005C, HHSN268201100006C, HHSN268201100007C, HHSN268201100008C, HHSN268201100009C, HHSN268201100010C, HHSN268201100011C, and HHSN268201100012C). Neurocognitive data in ARIC is collected by U01 2U01HL096812, 2U01HL096814, 2U01HL096899, 2U01HL096902, 2U01HL096917 from the NIH (NHLBI, NINDS, NIA and NIDCD), and with previous brain MRI examinations funded by R01-HL70825 from the NHLBI. CHS research was supported by contracts HHSN268201200036C, HHSN268200800007C, N01HC55222, N01HC85079, N01HC85080, N01HC85081, N01HC85082, N01HC85083, N01HC85086, and grants U01HL080295 and U01HL130114 from the NHLBI with additional contribution from the National Institute of Neurological Disorders and Stroke (NINDS). Additional support was provided by R01AG023629, R01AG15928, and R01AG20098 from the NIA. FHS research is supported by NHLBI contracts N01-HC-25195 and HHSN268201500001I. This study was also supported by additional grants from the NIA (R01s AG054076, AG049607 and AG033040 and NINDS (R01 NS017950). The ERF study as a part of EUROSPAN (European Special Populations Research Network) was supported by European Commission FP6 STRP grant number 018947 (LSHG-CT-2006-01947) and also received funding from the European Community’s Seventh Framework Programme (FP7/2007-2013)/grant agreement HEALTH-F4- 2007-201413 by the European Commission under the programme “Quality of Life and Management of the Living Resources” of 5th Framework Programme (no. QLG2-CT-2002- 01254). High-throughput analysis of the ERF data was supported by a joint grant from the Netherlands Organization for Scientific Research and the Russian Foundation for Basic Research (NWO-RFBR 047.017.043). The Rotterdam Study is funded by Erasmus Medical Center and Erasmus University, Rotterdam, the Netherlands Organization for Health Research and Development (ZonMw), the Research Institute for Diseases in the Elderly (RIDE), the Ministry of Education, Culture and Science, the Ministry for Health, Welfare and Sports, the European Commission (DG XII), and the municipality of Rotterdam. Genetic data sets are also supported by the Netherlands Organization of Scientific Research NWO Investments (175.010.2005.011, 911-03-012), the Genetic Laboratory of the Department of Internal Medicine, Erasmus MC, the Research Institute for Diseases in the Elderly (014-93-015; RIDE2), and the Netherlands Genomics Initiative (NGI)/Netherlands Organization for Scientific Research (NWO) Netherlands Consortium for Healthy Aging (NCHA), project 050-060-810. All studies are grateful to their participants, faculty and staff. The content of these manuscripts is solely the responsibility of the authors and does not necessarily represent the official views of the National Institutes of Health or the U.S. Department of Health and Human Services.

The FUS cohorts include: the Alzheimer’s Disease Research Centers (ADRC) (P30 AG062429, P30 AG066468, P30 AG062421, P30 AG066509, P30 AG066514, P30 AG066530, P30 AG066507, P30 AG066444, P30 AG066518, P30 AG066512, P30 AG066462, P30 AG072979, P30 AG072972, P30 AG072976, P30 AG072975, P30 AG072978, P30 AG072977, P30 AG066519, P30 AG062677, P30 AG079280, P30 AG062422, P30 AG066511, P30 AG072946, P30 AG062715, P30 AG072973, P30 AG066506, P30 AG066508, P30 AG066515, P30 AG072947, P30 AG072931, P30 AG066546, P20 AG068024, P20 AG068053, P20 AG068077, P20 AG068082, P30 AG072958, P30 AG072959), Alzheimer’s Disease Neuroimaging Initiative (ADNI) (U19AG024904), Amish Protective Variant Study (RF1AG058066), Cache County Study (R01AG11380, R01AG031272, R01AG21136, RF1AG054052), Case Western Reserve University Brain Bank (CWRUBB) (P50AG008012), Case Western Reserve University Rapid Decline (CWRURD) (RF1AG058267, NU38CK000480), CubanAmerican Alzheimer’s Disease Initiative (CuAADI) (3U01AG052410), Estudio Familiar de Influencia Genetica en Alzheimer (EFIGA) (5R37AG015473, RF1AG015473, R56AG051876), Genetic and Environmental Risk Factors for Alzheimer Disease Among African Americans Study (GenerAAtions) (2R01AG09029, R01AG025259, 2R01AG048927), Gwangju Alzheimer and Related Dementias Study (GARD) (U01AG062602), Hillblom Aging Network (2014-A-004-NET, R01AG032289, R01AG048234), Hussman Institute for Human Genomics Brain Bank (HIHGBB) (R01AG027944, Alzheimer’s Association “Identification of Rare Variants in Alzheimer Disease”), Ibadan Study of Aging (IBADAN) (5R01AG009956), Longevity Genes Project (LGP) and LonGenity (R01AG042188, R01AG044829, R01AG046949, R01AG057909, R01AG061155, P30AG038072), Mexican Health and Aging Study (MHAS) (R01AG018016), Multi-Institutional Research in Alzheimer’s Genetic Epidemiology (MIRAGE) (2R01AG09029, R01AG025259, 2R01AG048927), Northern Manhattan Study (NOMAS) (R01NS29993), Peru Alzheimer’s Disease Initiative (PeADI) (RF1AG054074), Puerto Rican 1066 (PR1066) (Wellcome Trust (GR066133/GR080002), European Research Council (340755)), Puerto Rican Alzheimer Disease Initiative (PRADI) (RF1AG054074), Reasons for Geographic and Racial Differences in Stroke (REGARDS) (U01NS041588), Research in African American Alzheimer Disease Initiative (REAAADI) (U01AG052410), the Religious Orders Study (ROS) (P30 AG10161, P30 AG72975, R01 AG15819, R01 AG42210), the RUSH Memory and Aging Project (MAP) (R01 AG017917, R01 AG42210Stanford Extreme Phenotypes in AD (R01AG060747), University of Miami Brain Endowment Bank (MBB), University of Miami/Case Western/North Carolina A&T African American (UM/CASE/NCAT) (U01AG052410, R01AG028786), and Wisconsin Registry for Alzheimer’s Prevention (WRAP) (R01AG027161 and R01AG054047).

The four LSACs are: the Human Genome Sequencing Center at the Baylor College of Medicine (U54 HG003273), the Broad Institute Genome Center (U54HG003067), The American Genome Center at the Uniformed Services University of the Health Sciences (U01AG057659), and the Washington University Genome Institute (U54HG003079). Genotyping and sequencing for the ADSP FUS is also conducted at John P. Hussman Institute for Human Genomics (HIHG) Center for Genome Technology (CGT).

Biological samples and associated phenotypic data used in primary data analyses were stored at Study Investigators institutions, and at the National Centralized Repository for Alzheimer’s Disease and Related Dementias (NCRAD, U24AG021886) at Indiana University funded by NIA. Associated Phenotypic Data used in primary and secondary data analyses were provided by Study Investigators, the NIA funded Alzheimer’s Disease Centers (ADCs), and the National Alzheimer’s Coordinating Center (NACC, U24AG072122) and the National Institute on Aging Genetics of Alzheimer’s Disease Data Storage Site (NIAGADS, U24AG041689) at the University of Pennsylvania, funded by NIA. Harmonized phenotypes were provided by the ADSP Phenotype Harmonization Consortium (ADSP-PHC), funded by NIA (U24 AG074855, U01 AG068057 and R01 AG059716) and Ultrascale Machine Learning to Empower Discovery in Alzheimer’s Disease Biobanks (AI4AD, U01 AG068057). This research was supported in part by the Intramural Research Program of the National Institutes of health, National Library of Medicine. Contributors to the Genetic Analysis Data included Study Investigators on projects that were individually funded by NIA, and other NIH institutes, and by private U.S. organizations, or foreign governmental or nongovernmental organizations.

An up to date acknowledgment statement can be found on the ADSP site: <https://www.niagads.org/adsp/content/acknowledgement-statement>.

**eMethods.** Supplementary Note

**Enrollment criteria**

Stanford’s Iqbal Farrukh and Asad Jamal Alzheimer’s Disease Research Center (ADRC) is a cohort of healthy older controls and patients with AD and related neurological disorders (n=323 with LRS and short-read NGS, age range 45-92 years old, 169 females and 154 males, healthy controls = 150, mild cognitive impairment (MCI) individuals = 60, AD cases = 30, other diagnoses = 83). All participants underwent a history and neurological exam, cognitive testing, and blood draw. Most participants also underwent brain imaging including MRI and amyloid PET scanning. Roughly 1/3 of the participants also provided cerebrospinal fluid (CSF). Diagnoses were determined in a consensus conference meeting comprised of neurologists and neuropsychologists using standard clinical criteria for AD, MCI, and related disorders such as Parkinson’s disease and Lewy body disease.

The Stanford Aging and Memory Study (SAMS) is a cohort of cognitively unimpaired older individuals (n=109 with LRS and short-read NGS, age range 60-88 years old, 58 females and 51 males). SAMS eligibility criteria include normal or corrected vision and hearing, native English speaking, no neurologic or psychiatric disease history, Clinical Dementia Rating score of zero, and normal performance on standardized neuropsychological testing. Participants underwent CSF collection, plasma collection, and brain imaging, including MRI and amyloid PET. Unimpaired cognitive status was confirmed in a consensus conference meeting comprised of neurologists and neuropsychologists using standard clinical criteria.

The Washington University St Louis (WUSTL) data enrolled participants as part of the Knight local ADRC (n=1,979 with short-read NGS, transcriptomics and/or CSF and/or plasma proteomics, age range 18-103 years old, 1034 females and 945 males, healthy controls = 1005, AD cases = 858, other diagnoses = 116). The WUSTL cohort includes longitudinally assessed community-dwelling adults older than 18 years old via prospective studies of memory and aging since 1979. All participants are required to participate in core study procedures, including annual longitudinal clinical assessments, neuropsychological testing, neuroimaging, and biofluid biomarker studies. Samples have been obtained from over 5,510 participants, including 2,426 AD cases, 694 AD-related dementia cases, 148 Frontotemporal dementia cases, 88 dementia with Lewy Body cases, and 2,156 cognitively normal healthy individuals.

**Multi-omics cohort aggregated at Washington University St Louis (WUSTL)**

Three omics modalities were analyzed as part of this study: plasma proteomics dataset, cerebrospinal fluid (CSF) proteomics dataset, and blood RNAseq dataset. Within each, participants considered for quantitative traits locus (QTL) analysis were required to have short-read NGS-based genotype data. Some analyses were restricted to healthy control-only participants, while others (when non-specified) considered all participants regardless of the disease status. Below we describe the acquisition protocol for each omics modality.

***CSF Proteomics Quality Control***

Proteomic data generation and QC were done as reported before^1,2^. Briefly, CSF samples were collected through lumbar puncture from participants after an overnight fast. Samples were processed and stored at -80 ⁰C until they were sent for protein measurement. In total, 3591 CSF samples were collected. CSF samples from Washington University were sent for protein measurement using the SOMAscan platform^3^ (SOMAscan7k), measuring 7596 aptamers. The SOMAscan7k data underwent initial normalization by SomaLogic. At the sample level, they performed hybridization normalization. The aptamers were then divided into S1, S2, and S3 normalization groups based on signal-to-noise ratio. SomaLogic then performed median normalization to remove biases due to protein concentration, pipetting variation, reagent concentration variation, and assay timing among others. Each sample was normalized to a reference to account for technical and biological variance. This step was performed with iterative Adaptive Normalization by Maximum Likelihood (ANML) until convergence was reached. This method is a modification of median normalization. Further quality control was performed on the normalized SOMAscan7k data provided by SomaLogic according to an in-house protocol for CSF samples. Aptamers were removed if they failed either of two criteria: first, if the maximum absolute difference between the aptamer scale factor and median scale factor of any plate is > 0.5; second, if the median cross-plate coefficient of variation (CV) was >0.15. Interquartile range (IQR) was then calculated for every aptamer based on log-10 transformed aptamer levels. Aptamer values outside of 1.5-fold of the IQR were replaced with NA values. Aptamers with call rate <65% (aptamer measurement in less than 65% of samples) were excluded, and the same criteria were used to remove samples. The call rate for aptamers was then recalculated and a more stringent call rate threshold of 85% was applied. The sample call rate was recalculated after aptamer removal and a call rate threshold of 85% was applied at the sample level. Overall, 1210 participants of European ancestry with short-read NGS passed QC and were included in this study.

***Plasma Proteomics Quality Control***

Plasma samples were collected through blood draws from participants at Knight ADRC from Washington University. Samples were processed and stored at -80 ⁰C until they were sent for protein measurement. 7548 aptamers were measured before proteomics QC with the SOMAscan platform^3^ (SOMAscan7k). The SOMAscan7k data were initially normalized by SomaLogic. At the sample level, they performed hybridization normalization. The aptamers were then divided into S1, S2, and S3 normalization groups based on signal-to-noise ratio. SomaLogic then performed median normalization to remove biases due to protein concentration, pipetting variation, reagent concentration variation, and assay timing among others. Each sample was normalized to a reference to account for technical and biological variance. This step was performed with iterative Adaptive Normalization by Maximum Likelihood (ANML) until convergence was reached. This method is a modification of median normalization. Plasma proteomics data were next QCed in-house with the 7 steps: First, we transformed the protein level into log-10 scale and filtered aptamers per limit of detection, scale factor difference, and coefficient of variation; Second, we used IQR-based outlier expression level to flag outliers; Third, we removed aptamers and samples with <65% call rate; Fourth, we re-calculated call rate for analytes and removed aptamers with call rate <85%; Fifth, we re-calculated missing rate for participants and remove participants with < 85% call rate cut-off; Sixth, we back transformed the protein-level into raw values; Seventh, we removed the non-human aptamers. Overall, 1150 participants of European ancestry and 200 participants of African ancestry with short-read NGS passed QC and were included in this study.

***Blood-based RNASeq (transcriptomics) Quality Control***

RNA-seq data was first processed with fastqc software. The data was next aligned to the GRCh38 reference genome using STAR (version 2.7.8a). To quantify the count per gene, Salmon (version 1.7.0) was used to infer gene expression for all samples to quantify the coding transcripts of Homo Sapiens reference genome (hg38) accessed from the GENCODE database. Overall, 428 participants of European ancestry with short-read NGS passed QC and were included in this study.

***Short-read Next Generation Sequencing (NGS) Genotype Quality Control***

We performed joint analysis and quality control (QC) for all samples. Whether we started from BAM or CRAM files, all were converted to fastq files. Alignment was conducted against GRCh38.p13 genome reference. Variant calling was performed for WGS following GATK’s 4.2 Best Practices. WGS data was filtered with allele-specific VQSR filtering with a truth sensitivity 99.7%. WGS data was filtered to remove low-complexity regions, and regions with excessive depth. Only those variants and indels that fell within the above 99.9% confidence threshold were considered for analysis; additional variant filters included allele-balance (AB = 0.3–0.7) and missingness (geno = 0.05). Variants out of Hardy Weinberg equilibrium (P<1x10^-8^) or with differential missingness between cases and controls, WGS, or different sequencing platforms were removed from the analysis. In addition, individuals with more than 2% of missing variants and whose genotype data indicated a sex discordant from the clinical database were removed from the dataset. The principal component analysis to account for population stratification was performed using plink1.9’s pca function. Overall, 5510 participants passed WGS QC.

**Validation of the 3’UTR insertion genotype with IGV for Stanford LRS participants**

The genotypes of rs1990622, rs3173615, and the 3’ UTR insertion were extracted for participants with both LRS and short-read NGS available. 18 individuals with discordant doses of the three variants in LRS – where the dose of any of the three variants differed from any other – were identified for validation with IGV. For these participants, LRS genome alignments were visualized in IGV^18^ and the dose of the *TMEM106B* 3’ UTR deletion was determined by the following criteria: (1) the dose was set to 0 if no reads contained the deletion, (2) the dose was set to 1 if at least two but not all reads contained the deletion, and (3) the dose was set to 2 if all reads contained the deletion. After the visualization of reads in IGV, 15 individuals among the 18 were actually concordant for the three genotypes, two individuals were discordant, and one was set as unknown for the deletion due to having only one read with the deletion among 10 reads overlapping this region. Regarding the two discordant individuals, one had the SV deletion genotype concordant with rs1990622 but discordant with rs3173615, while the other had the SV deletion genotype discordant with rs1990622 but concordant with rs3173615.

***TMEM106B* 3’UTR insertion genotype quality control in ADSP**

A *TMEM106B* 3’ UTR deletion was identified in Biograph and Manta SV calls (chr7:12242077; SVLEN=-322; SVTYPE=DEL, with Manta SVLEN varying between -321 and -323 base pairs for a few unique samples). 612 samples were duplicates of 288 unique participants. Among these duplicate individuals, 45 Biograph SV genotypes were discordant among replicates while 6 were discordant in Manta. Among these discordant genotypes, 5 were concordant in Manta and Biograph for a given sample/instance of one participant, albeit discordant in the other sample, and thus within participant discrepancy was related to the actual reads in the sample. These 5 participants were thus excluded from further analysis. The remaining discordant participant in Manta had 6 replicate samples, and the other 5 were concordant, thus the SV genotype was set to this majority. In all instances of Biograph discordances, the genotype of one of the replicates was concordant with the Manta genotypes. As such and upon manual inspection, Biograph calls were judged to be less reliable than Manta for this particular deletion, and analysis in ADSP refers to SV genotype by Manta. Among the 612 duplicate samples, all SNVs (rs1990622, and rs3173615) were concordant across duplicates. One instance among duplicated genotypes was kept for analysis leading to 16,582 unique participants in ADSP.

**WUSTL dataset genomics analysis**

### ***Plasma pQTL analysis***

A linear regression model was used from lm function in R for each protein. Protein abundances were log-10 transformed first and z-value normalization next. Covariates were age, sex, genotype PC1-10, and proteomics PC 1-2. The final sample size for EUR and AFR pQTL analyses were 1150 and 200. The sample sizes for healthy control only were 120 AFR and 711 EUR.

### ***CSF pQTL analysis***

A linear regression model was used from lm function in R for each protein. Protein-abundances were log-10 transformed first and z-value normalization next. Covariates were genotype PC1-10 and 60 PEER factors, which were highly associated with age, and sex. The final sample size for EUR pQTL analysis was 1210 and the sample size for healthy control-only pQTL analysis was 588.

### ***Blood eQTL analysis***

Linear regression was used from lm function in R. RNA *TMEM106B* gene expression levels were log10 transformed from the raw count values and covariates were age and sex. The sample size for healthy control only EUR eQTL is 428.

### ***Differential abundance analysis of the PGRN mutation carriers vs the non-carriers***

Linear regression was used from lm function in R for each protein. The plasma protein TMEM106B and GRN levels were log-10 transformed first and z-value normalization next. Significance (p-value on **Figure 3** boxplots) was assessed based on the estimated difference from the linear regression of mutation status on protein level after adjusting for age, sex, and proteomic PC1 and PC2.

***Differential abundance analysis of Alzheimer’s cases vs controls***

The same methods were used here as for the PRGN mutation carriers vs non-carriers described above. Analyses were done both on plasma and cerebrospinal fluid protein levels of TMEM106B and GRN.

**eFigure 1. Effect of age on TMEM106B protein level in CSF and plasma.**

A) Scatterplot of normalized plasma TMEM106B protein level against age at plasma draw. The blue line is added for linear smoothing with gray area as the confidence interval.

B) Scatterplot of normalized CSF TMEM106B protein level against the age at CSF draw. The blue line is added for linear smoothing with gray area as the confidence interval.

C) Table summarizing the correlation analyses of age and TMEM10B protein levels in CSF and plasma.

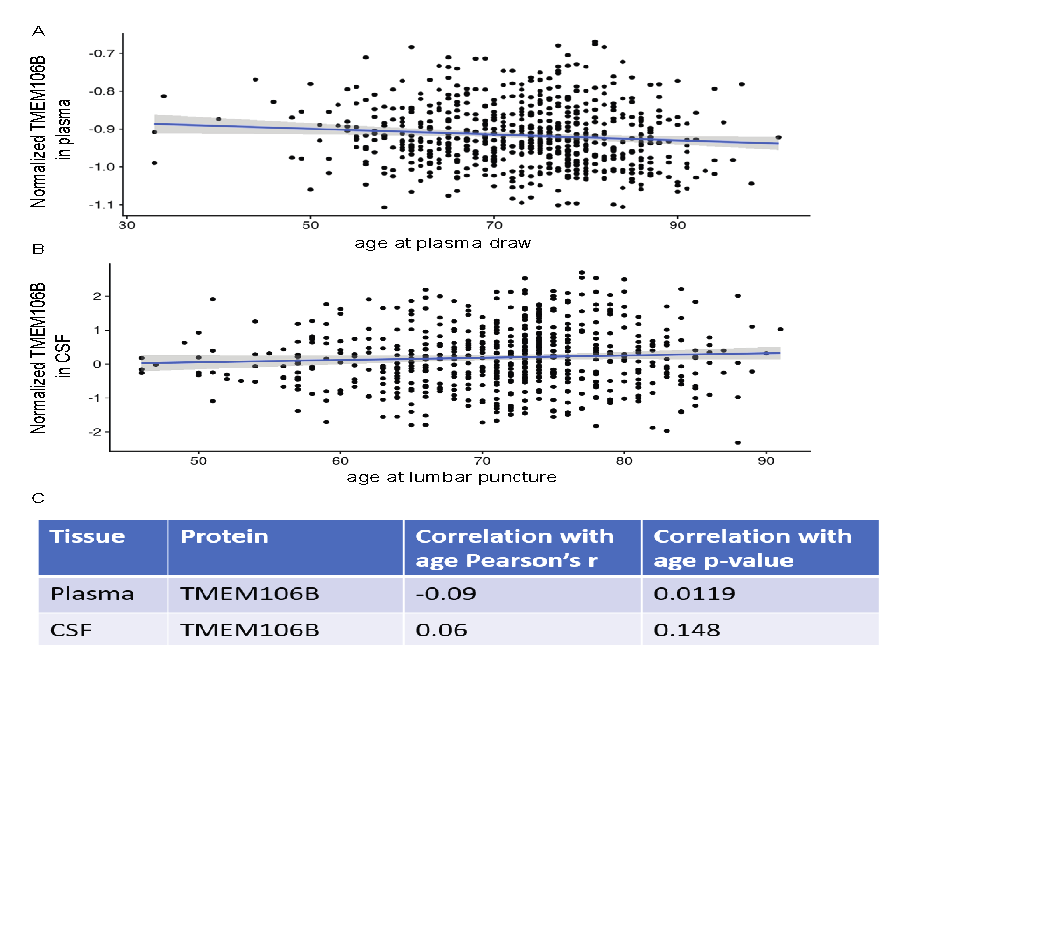

**eFigure 2. Effect of age on TMEM106B or GRN levels in two tissue types in two ancestries**

A) Scatterplot of age effects on normalized plasma TMEM106B in AFR ancestry. The blue line is added for linear smoothing with gray area as confidence interval.

B) Scatterplot of age effects on normalized plasma TMEM106B in EUR ancestry. The blue line is added for linear smoothing with gray area as confidence interval.

C) Scatterplot of age effects on normalized CSF TMEM106B in EUR ancestry. The blue line is added for linear smoothing with gray area as confidence interval.

D) A table summarizing the correlation coefficients and p-value for the panels A-C.

E) Scatterplot of age effects on normalized plasma GRN in AFR ancestry. The blue line is added for linear smoothing with gray area as confidence interval.

F) Scatterplot of age effects on normalized plasma GRN in EUR ancestry. The blue line is added for linear smoothing with gray area as confidence interval.

G) Scatterplot of age effects on normalized CSF GRN in EUR ancestry. The blue line is added for linear smoothing with gray area as confidence interval.

H) A table summarizing the correlation coefficients and p-value for the panels E-G.

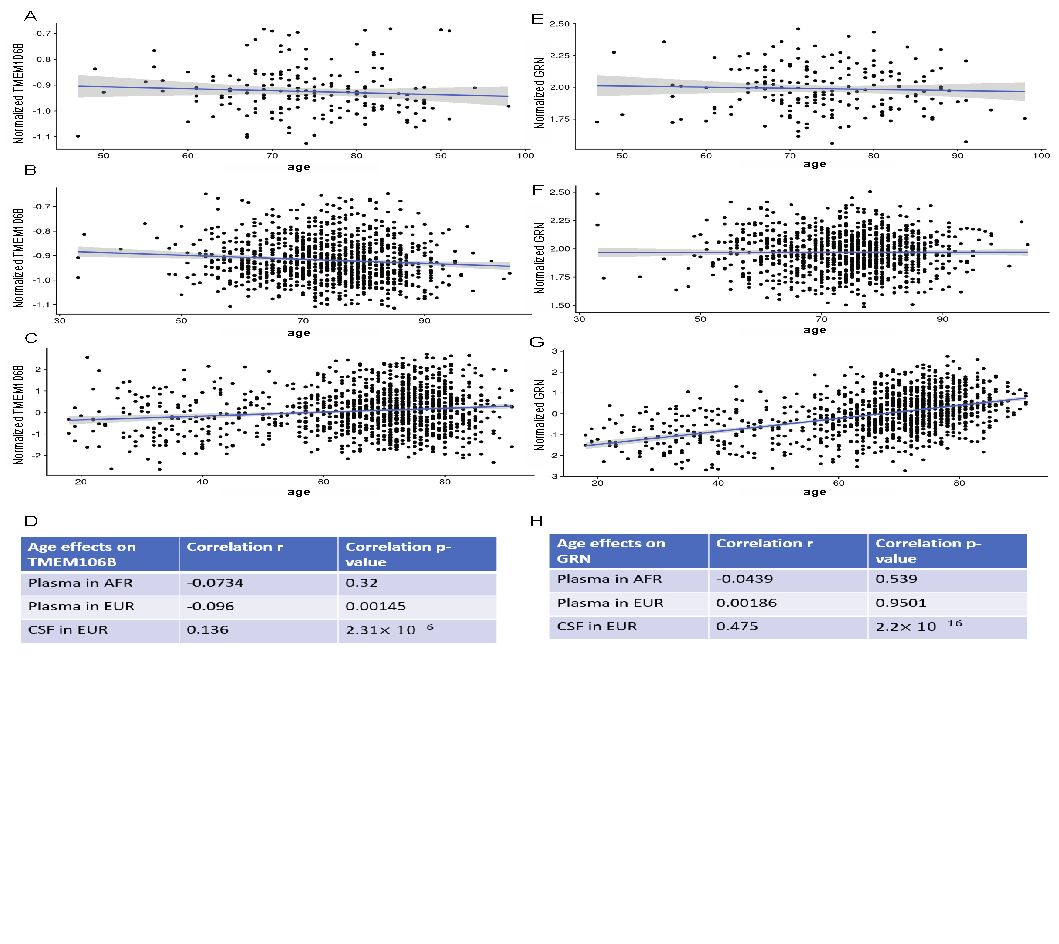

**eFigure 3. Effect of sex on TMEM106B or GRN levels in two tissue types in two ancestries.**

A) Boxplot of sex effects on normalized plasma TMEM106B in AFR ancestry.

B) Boxplot of sex effects on normalized plasma TMEM106B in EUR ancestry.

C) Boxplot of sex effects on normalized CSF TMEM106B in EUR ancestry.

D) Table summarizing the sex difference and p-value from the linear regression for panels A-C.

E) Boxplot of sex effects on normalized plasma GRN in AFR ancestry.

F) Boxplot of sex effects on normalized plasma GRN in EUR ancestry.

G) Boxplot of sex effects on normalized CSF GRN in EUR ancestry.

H) Table summarizing the sex difference and p-value from the linear regression for panels E-G.

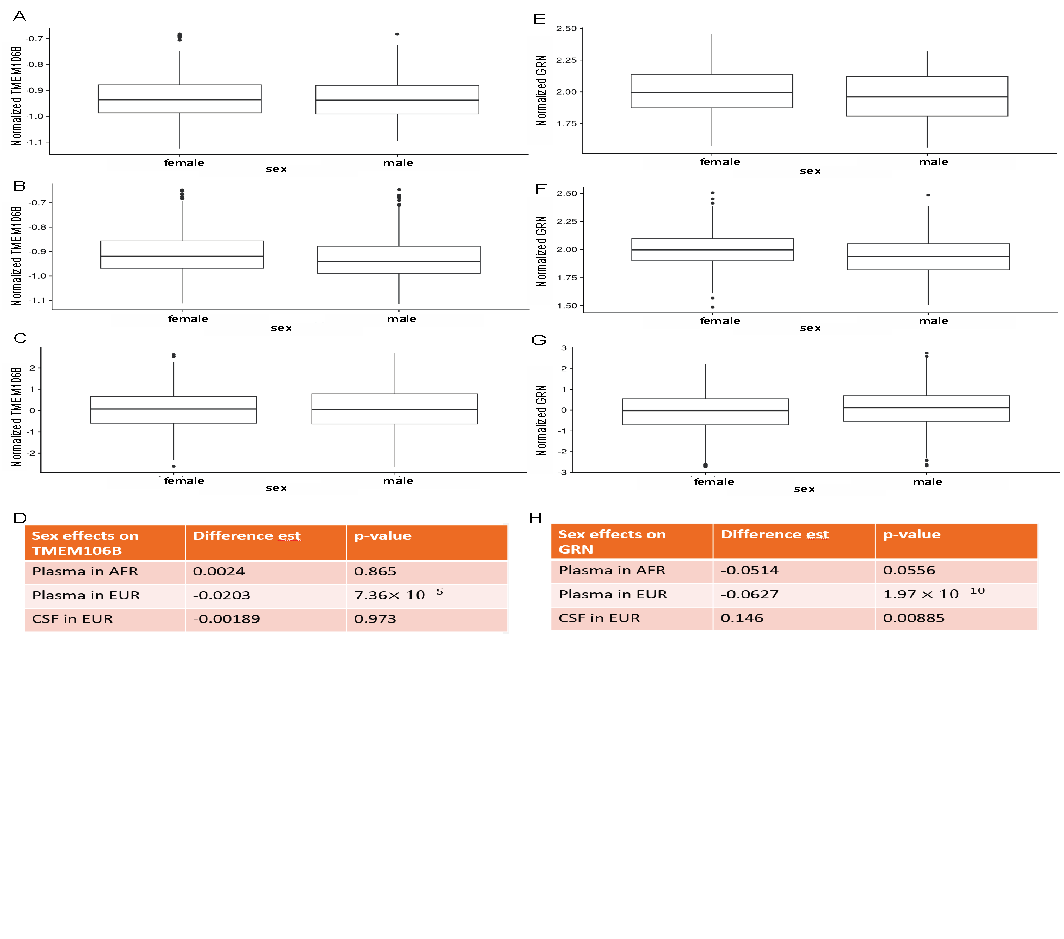

**eFigure 4**. **Colocalization analyses between the AD and FTLD-TDP GWAS and the plasma pQTL GWAS (deCODE and ARIC).** Note that the FTLD-TDP GWAS only included directly genotyped variants, i.e., not imputed, and thus the intersection of this GWAS and the pQTL GWAS (bottom panels) led to a small number of variants (300 to 600) compared to the intersection of the AD GWAS and the pQTL GWAS. As such, the colocalization analysis is less robust for the FTLD-TDP GWAS. For this reason, the bottom right panel, while indicative of a shared association does show PP3 higher than PP4 when formally computed (green variants spread across the diagonal). Note also, that the linkage (r^2^) is shown for the EUR 1000 genomes panel and thus does not apply well to the ARIC AA GWAS, which we still show for illustration purposes.

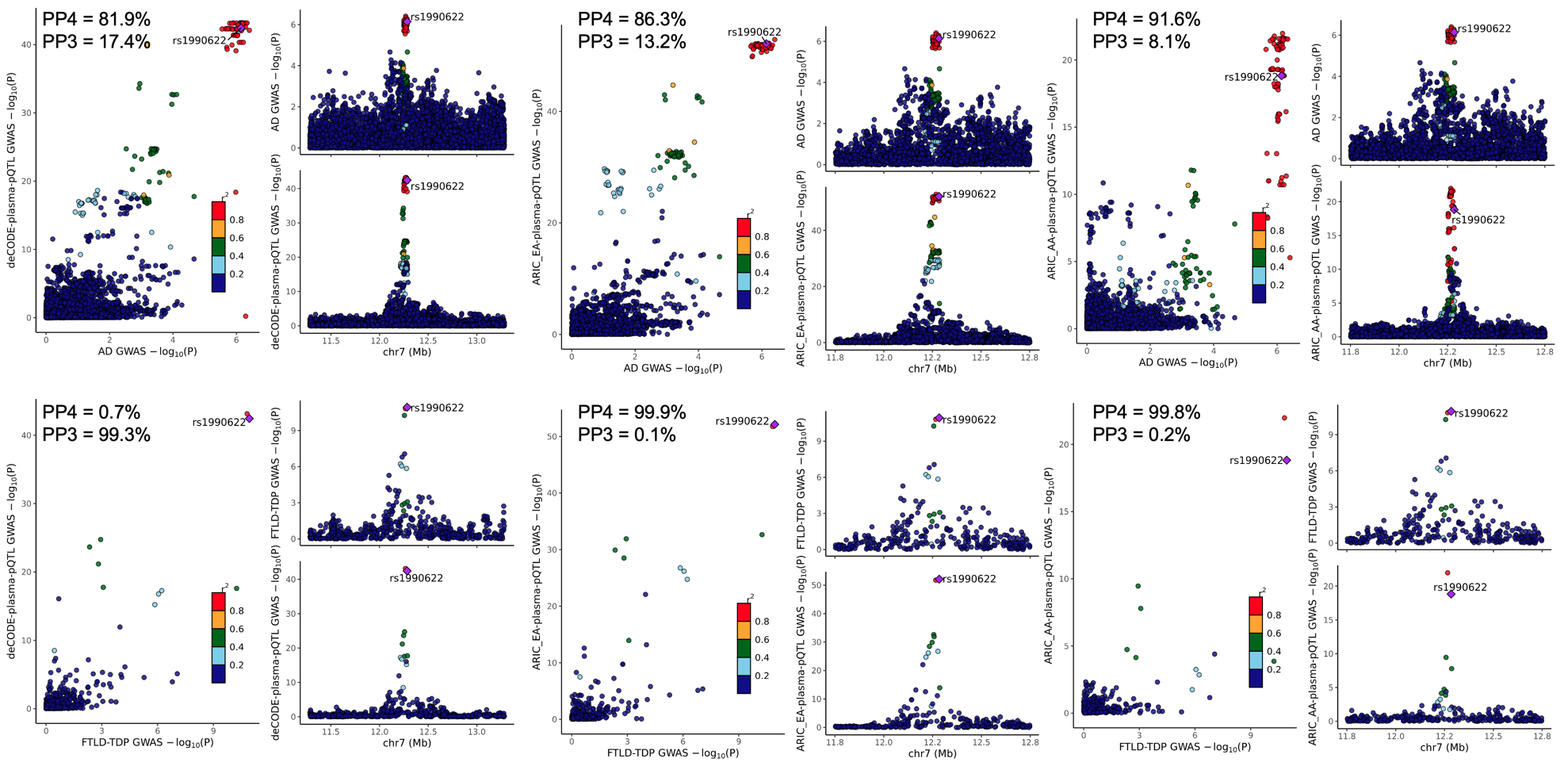

**eFigure 5.** **Effects of variants rs1990622(A) and rs3173615(C) on levels of protein TMEM106B in two tissues.**

A) Boxplot of normalized protein TMEM106B from EUR plasma samples across the genotype of rs3173615 (left) and rs1990622 (right) in healthy controls.

B) Boxplot of normalized protein TMEM106B from AFR plasma samples across the genotype of rs3173615 (left) and rs1990622 (right) in healthy controls.

C) Boxplot of normalized protein TMEM106B from EUR CSF samples across the genotype of rs3173615 (left) and rs1990622 (right) in healthy controls.

A summary of these results is reported in **eTable 2**.

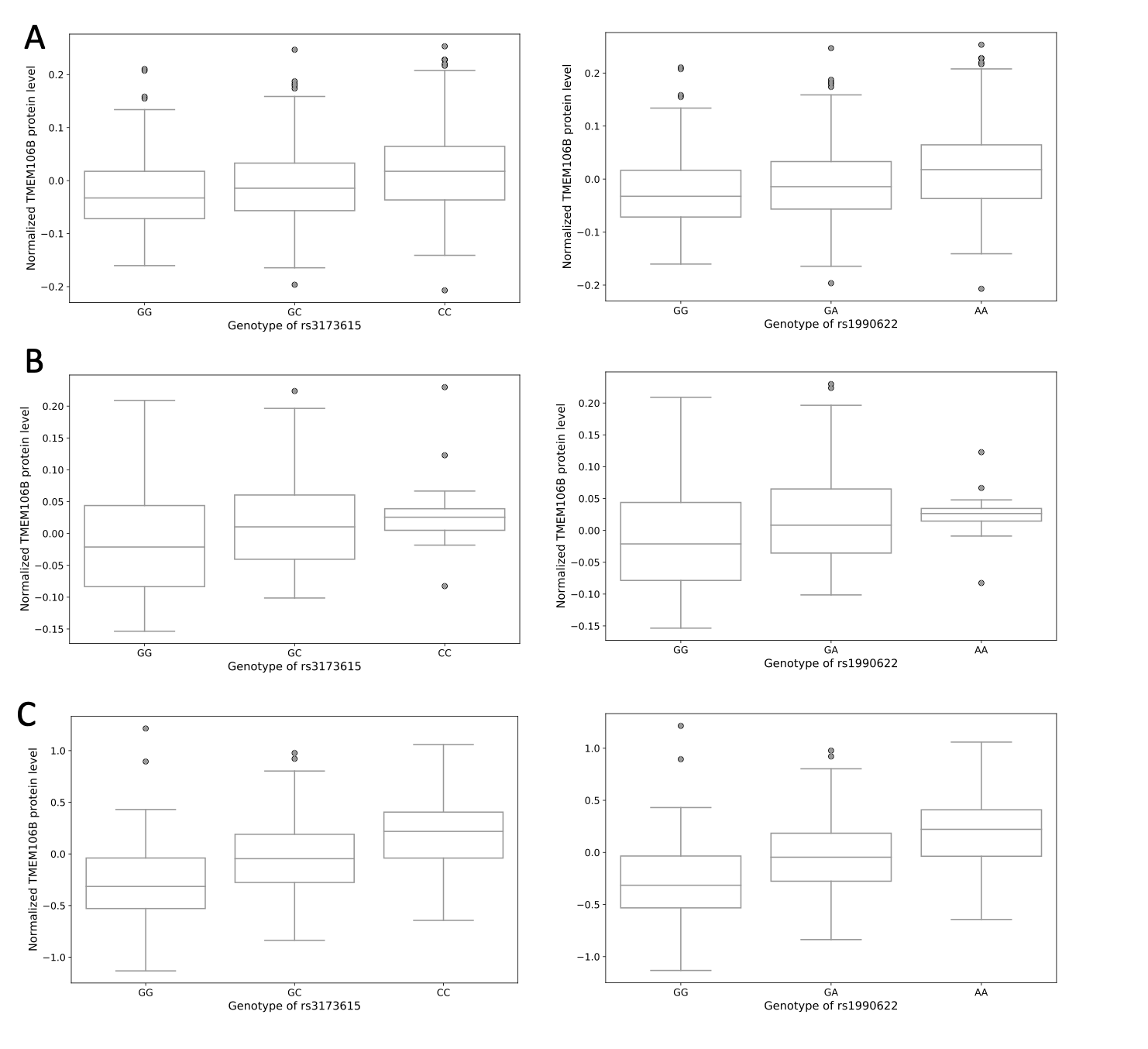

**eTable 1. Demographics per ‘omics modalities in the WUSTL Neurogenomics dataset.** (Age at blood draw, Sex, diagnostic status (AD: Alzheimer’s disease, HC: Healthy Control, Other: Other diagnosis).

| **Tissue** | **OmicsType** | **Ancestry** | **Age Range** | **Sex (female/male)** | **Status**  **(AD/HC/Other)** |
| --- | --- | --- | --- | --- | --- |
| CSF | Proteomics | EUR | 18-91 | 599/611 | 555/588/67 |
| Plasma | Proteomics | EUR | 33-104 | 617/533 | 367/711/72 |
| Plasma | Proteomics | AFR | 47-98 | 131/69 | 77/120/3 |
| Blood | RNAseq | EUR | 45-94 | 238/190 | 0/428/0 |

**eTable 2. Association of *TMEM106B* variants across tissues and in multiple populations.**

| **Tissue** | **SNP** | **Ancestry** | **Status** | **Effect size** | **pval** |
| --- | --- | --- | --- | --- | --- |
| Plasma | rs3173615 | EUR | Control | 0.0226 | 6.3×10^-8^ |
| Plasma | rs1990622 | EUR | Control | 0.0227 | 5.6×10^-8^ |
| Plasma | rs3173615 | AFR | Control | 0.0290 | 0.0321 |
| Plasma | rs1990622 | AFR | Control | 0.0290 | 0.0387 |
| CSF | rs3173615 | EUR | Control | 0.2800 | 7.2×10^-27^ |
| CSF | rs1990622 | EUR | Control | 0.2800 | 3.9×10^-27^ |

**eReferences.**

1.	Timsina J, Gomez-Fonseca D, Wang L, et al. Comparative Analysis of Alzheimer’s Disease Cerebrospinal Fluid Biomarkers Measurement by Multiplex SOMAscan Platform and Immunoassay-Based Approach 1. *Journal of Alzheimer’s Disease*. 2022;89(1):193-207. doi:10.3233/JAD-220399

2.	Cruchaga C, Western D, Timsina J, et al. Proteogenomic analysis of human cerebrospinal fluid identifies neurologically relevant regulation and informs causal proteins for Alzheimer’s disease. *researchsquare*. Published online June 9, 2023. https://www.researchsquare.com/article/rs-2814616/v1

3.	Gold L, Ayers D, Bertino J, et al. Aptamer-Based Multiplexed Proteomic Technology for Biomarker Discovery. *PLOS ONE*. 2010;5(12):e15004. doi:10.1371/journal.pone.0015004
